# Perception and Attitudes of Medical Students Regarding Postoperative Pain Management

**DOI:** 10.1101/2021.05.11.21257071

**Authors:** Fatema Johora, Asma Akter Abbasy, Fatiha Tasmin Jeenia, Ferdaush Ahmed Sojib, Mohsena Aktar, Priyanka Moitra, Jannatul Ferdoush, Afroza Momen, Mohammad Ali

## Abstract

**Background:** Postoperative pain remains a challenging problem, which requires a dynamic approach using a variety of treatment modalities to obtain an optimal outcome with respect to enhancing patient comfort and facilitating the recovery process. Multimodal analgesia signifies an increasingly popular approach to prevent postoperative pain. The approach includes administering a combination of opioid and nonopioid analgesics that act at different sites within the central and peripheral nervous systems in an effort to improve pain control while eliminating opioid-related adverse effects.

**Materials and Methods:** To understand the perception and attitude of 4^th^ year medical students, a structured questionnaire survey was done among six different medical colleges including government (Armed Forces Medical College, Cumilla Medical College and Colonel Malek Medical College, Manikganj) and non-government medical colleges (Army Medical College Bogura, Brahmanbaria Medical College and Chattogram International Medical College) of Bangladesh in October 2019. Total 340 students participated in the study.

**Result:** Among the 340 students, almost 336 (98.2%) students were agreed that postoperative pain management is an essential element of patient care. Only 4 (1.18%) respondents disagreed. On the other hand, regarding its influence on early recovery and reduction of hospital staying, 311 (91.47%) respondents agreed and 29 (8.53%) respondents were found as disagreed. Regarding the issue of opioid commonly induced respiratory depression, 257 (75.59%) respondents thought it is a common adverse event and 81 (23.82%) respondents disagreed. 206 (60.59%) respondents didn’t agree that opioid always produce addiction or tolerance and 134 (39.71%) respondents disagreed with the same issue. 294 (86) respondents agreed that multimodal analgesia increases patient’s cost and 46 (14%) disagreed. 249 (74%) students agreed that regional techniques are useful for postoperative pain management and 89 (26%) respondents disagreed.

**Conclusion:** Adequate post-operative pain management knowledge is necessary to reduce post-operative complications.

## Introduction

More than 230 million people undergo surgical procedure each year worldwide^[1]^ and more than 80% of patients experience acute postoperative pain ranging from moderate to severe pain.^[2-7]^ Effective postoperative pain control is essential component for the care of surgical patient. The advantages of effective postoperative analgesia includes patient comfort and therefore satisfaction, earlier mobilization, fewer pulmonary and cardiac complications, a reduced risk of deep vein thrombosis, reduction of postoperative nausea and delirium, faster recovery with less likelihood to development of neuropathic pain, and reduced healthcare cost. Inadequate pain control, apart from being inhumane, may result in increased morbidity and mortality because of acute organ dysfunction.^[8-11]^ Evidence suggests that inadequately controlled pain deleteriously affects quality of life, and functional recovery, the risk of postsurgical complications as well as the risk of persistent chronic pain.^[12-16]^ Insufficient patient education, age, sex, preoperative pain, type of surgery, incision size, psychological factors, preoperative anxiety, fear of complications associated with analgesic drugs, poor pain assessment and inadequate staffing found to be linked with severity of postoperative pain.^[8],[17-20]^ Inadequate postoperative pain assessment, absence of pain management guideline and giving least priority to postoperative pain are identified as major barriers to optimal post-operative analgesia.^[21]^ There are growing body of evidence suggesting that lack of proper knowledge and positive attitude of healthcare professionals towards postoperative pain management act as a potential barrier for adequate analgesia because of poor or absence of institutional training for pain management.^[22-26]^ Hence, the present study was carried out with the attempt to explore the perception and attitudes of future physicians regarding postoperative pain management as well as to create an awareness which might be insightful to achieve better patient care in future.

## Materials and Methods

### Place and duration of the study

This was a cross-sectional, structured questionnaire survey, conducted in six medical colleges of Bangladesh including government (Armed Forces Medical College, Cumilla Medical College and Colonel Malek Medical College, Manikganj) and non-government (Army Medical College Bogura, Brahmanbaria Medical College and Chattagram International Medical College) medical colleges in October 2019. Permission was taken from college authorities and informed consent was taken from the participants of the structured questionnaire survey.

### Procedure

The questionnaire survey was conducted among the 4^th^ year MBBS students of respective medical colleges. Researchers explain the nature and purpose of the survey to the students. The questionnaire was circulated among them who agreed to participate the study. This self-administered questionnaires were handed by the researchers to the students. To assure the quality, students filled and returned the questionnaire quickly during end of a lecture class. Consultation of medical textbooks, internet or discussion among themselves were not allowed. Participants who did not returned questionnaire in time were approached again. In case of losing the questionnaire, new questionnaire was provided to them. Students who did not return back their questionnaire after three approaches were considered as non-respondent. Questionnaire was validated before survey. As English is the official language of medical education in Bangladesh, questionnaire was in English.Total 340 students were participated in the study. To maintain confidentiality, responses were anyoymus. No demographic data was collected fromrespondents.Data was compiled, presented and appropriate statistical test was done in this study for drawing an appropriate conclusion.

## Result

**Figure 1** showed that 336 (98.2%) out of 340 students were agreed that postoperative pain management is an essential element of patient care. On the other hand, regarding its influence on early recovery and reduction of hospital staying, 29 (8.53%) respondents were found as disagreed. 257 (75.59%) students think that opioid commonly induces respiratory depression, and 206 (60.29%) students did not think that opioid always produce addiction (**Figure 2**).

**Figure 1:**
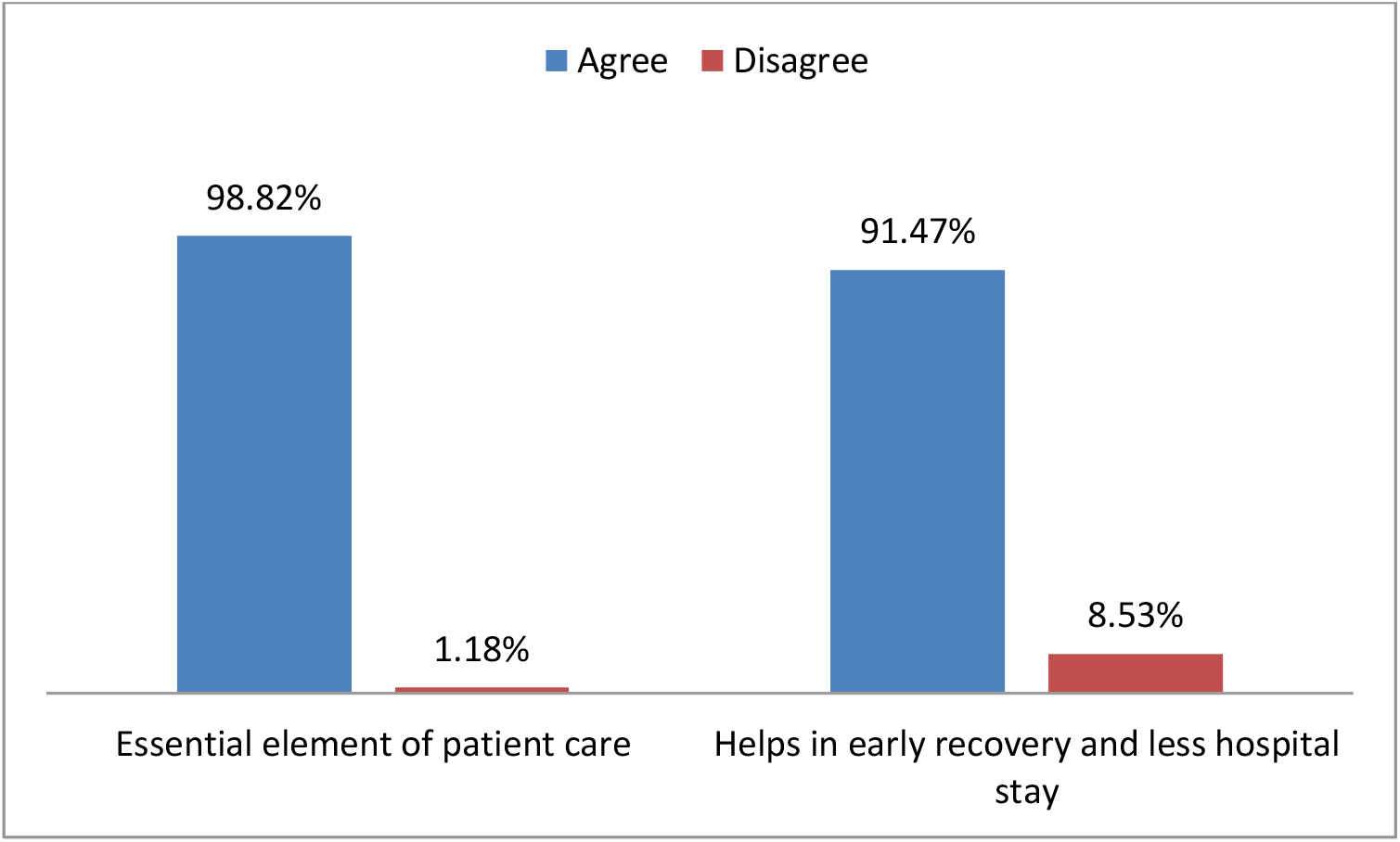
Perception and attitudes regarding importance of postoperative pain management.

**Figure 2:**
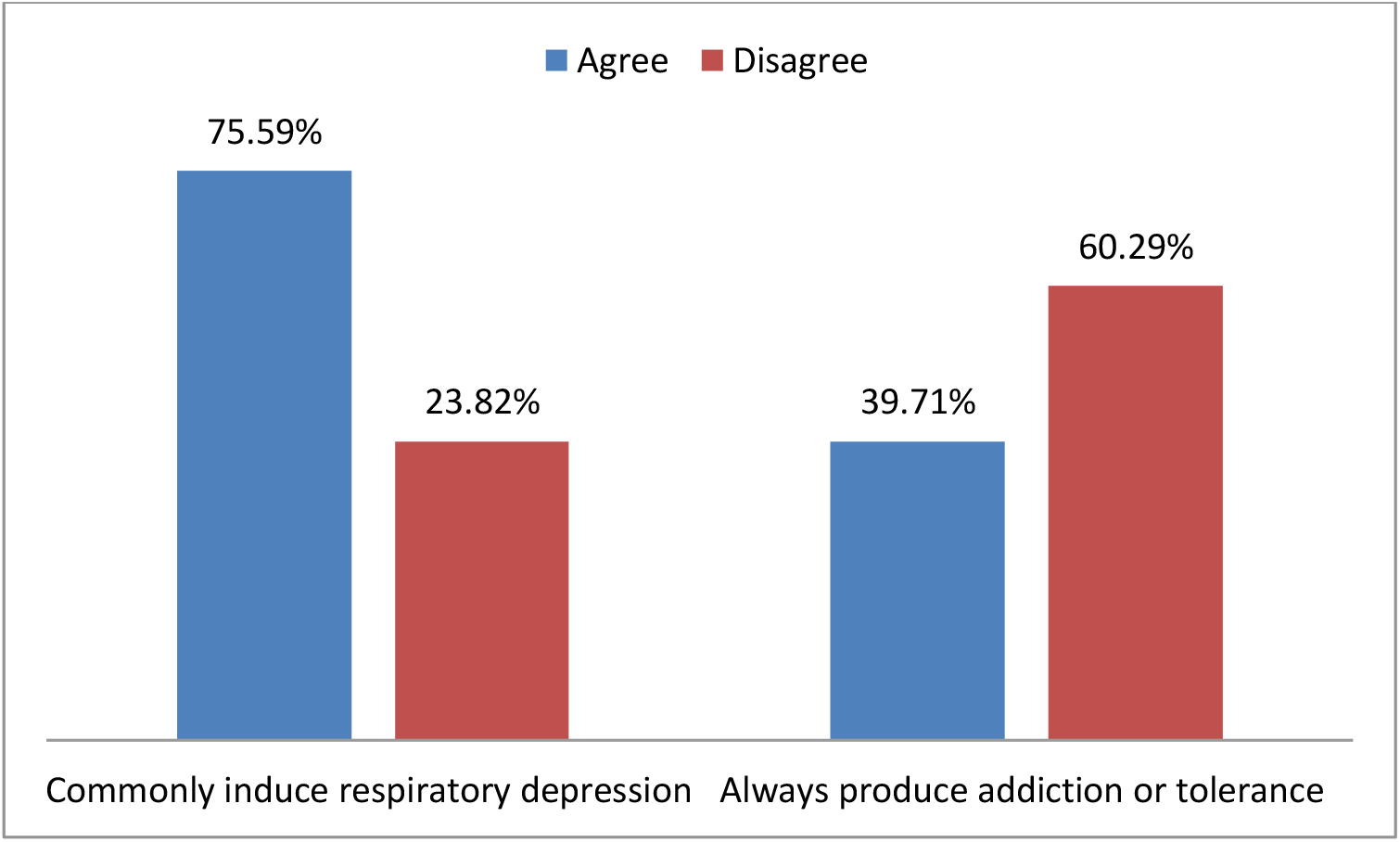
Perception and attitudes regarding opioid analgesic.

**Figure 3** showed that 294 (86%) students considered that use of multimodal analgesia increases patient’s cost. 249 (74%) students agreed that regional techniques are useful for postoperative pain management (**Figure 4**).

**Figure 3:**
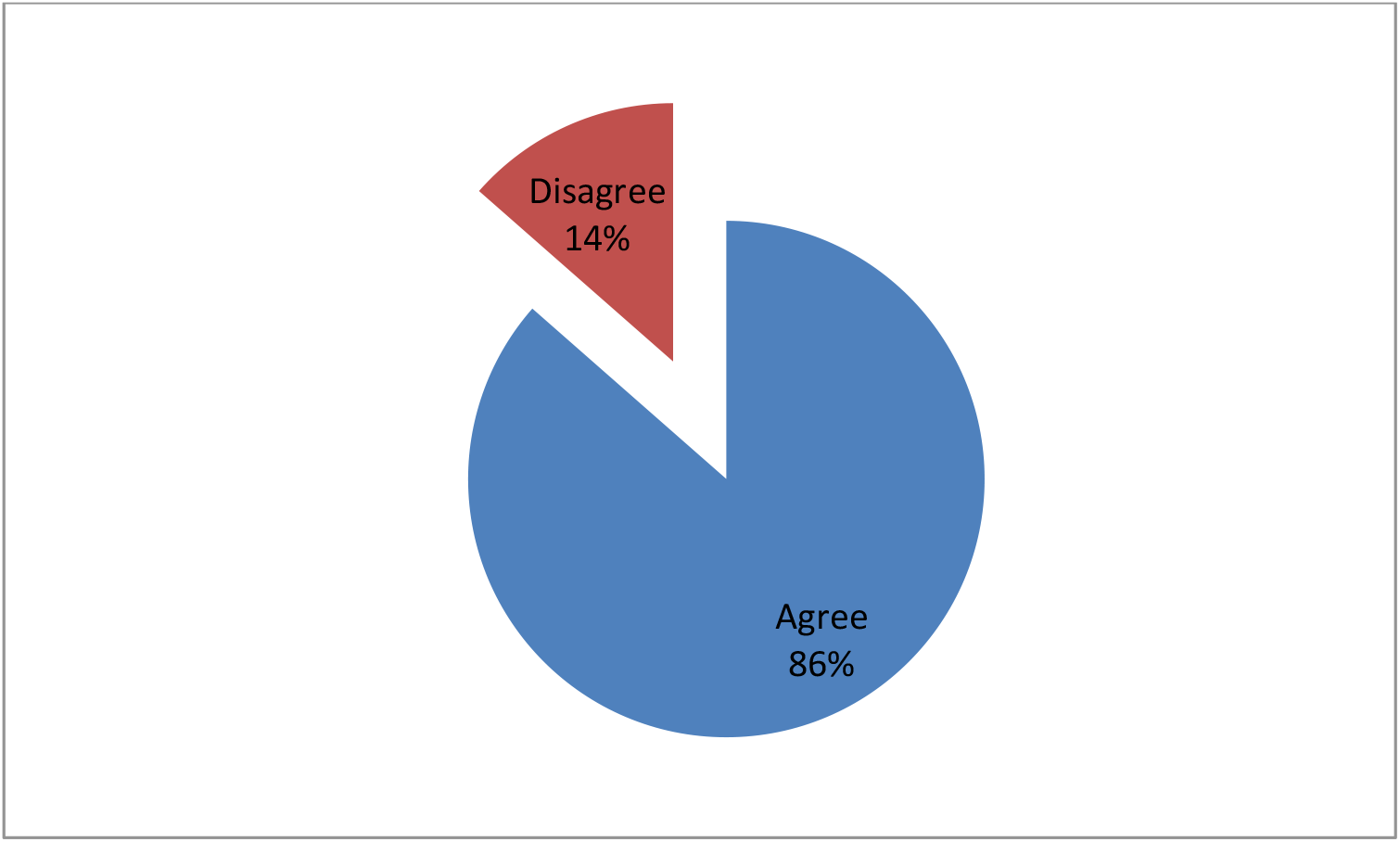
Perception towards multimodal analgesia.

**Figure 4:**
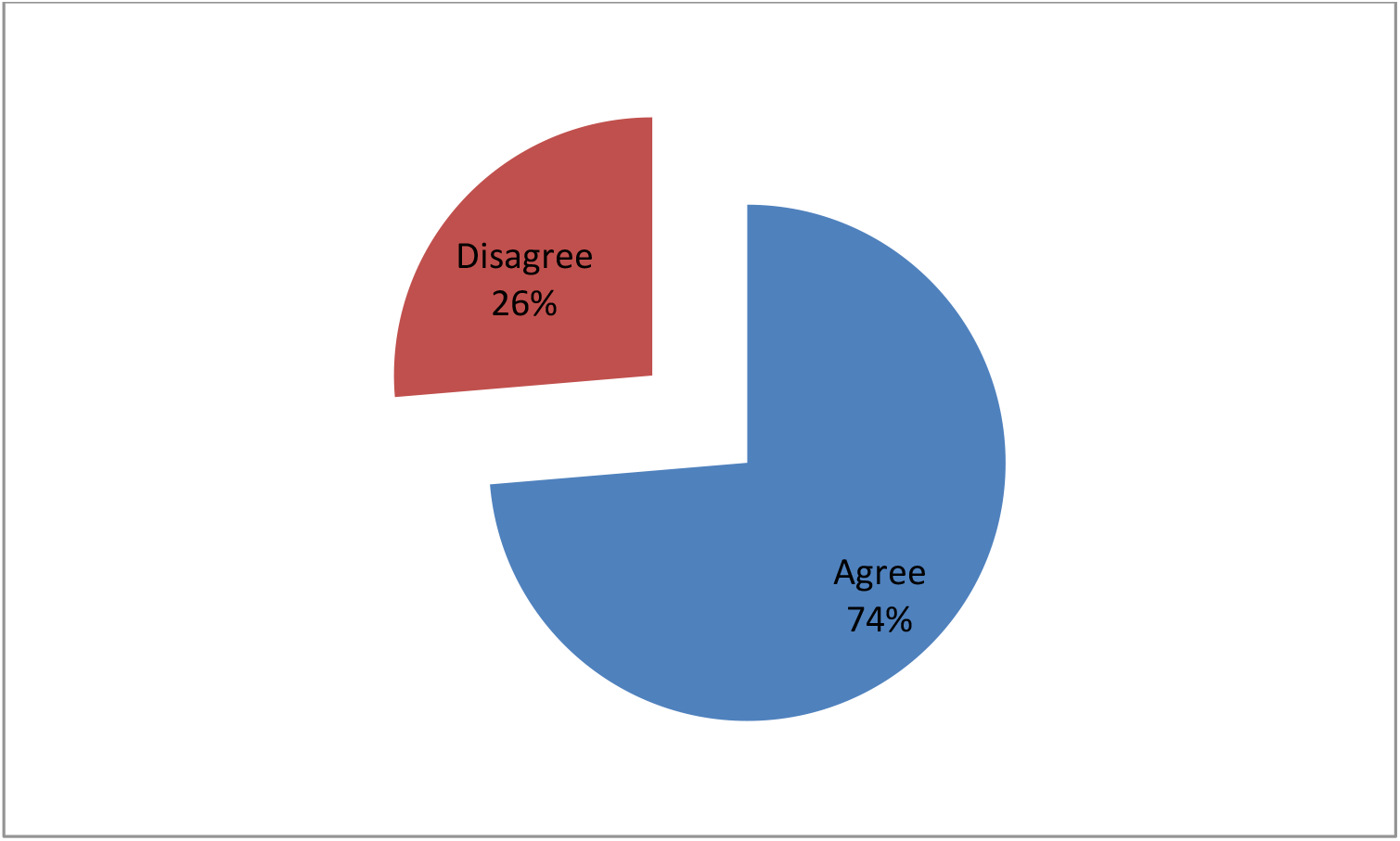
Perception towards regional techniques.

## Discussion

Provision of high-quality postoperative analgesia has become recognized as an important perioperative goal. Many preoperative, intraoperative and postoperative interventions and management strategies are available, and continue to evolve for reducing and managing postoperative pain that includes preoperative pain education, perioperative pain management planning, use of different pharmacological and non-pharmacological modalities. Despite of these efforts, knowlegde deficits and poor attitudes of physicians towards postoperative pain management stil now considered as an important barrier of surgical patient care both in developed and developing countries. There is no data available in this aspect among the physicians of Bangladesh. Current study was conducted in this context to assess the knowlege and perception of future physicians with an aim to develop an educational intervention in future for optimal patient care.

Effective control of acute postsurgical pain is essential for the patient not only in the short term but also in the long term to prevent the development of chronic pain, which can occur if early acute pain is prolonged. Clinical, psychological and institutional consequences may arise from inadequate pain management.^[2**6**]^ Postoperative pulmonary complications are common and a major contributor to the overall risk of surgery. It is associated with increased morbidity and mortality and adversely affects financial outcomes in health care. A multifaceted approach is necessary to reduce the incidence of postoperative complications ^[2**7**]^. In this study, most of the respondents had positive attitudes towards importance of postoperative pain management and this corresponds with similar studies conducted on practicing physicians.^**[2**3, 28-29]^

Opioid analgesics continue to play an important role in the acute treatment of moderate-to severe pain in the early postoperative period. When the most important source of nociceptive stimuli is visceral pain, good results may be achieved by opioids. Several opiod congenerse.g. morphine, fentanyl, sufentanil, buprenorphine, tramadol have been used.^[**30**]^ Although having excellent clinical efficacy, plethora of myths about opioid medication continue to pose as main reason for less use in clinical practice.Many physicians and patients hold exaggerated fear that addiction will result from opiod use during postoperative period^[**31-32**]^. In this study, 75.59% agreed about opiod induced respiratory depression, which corresponds with result found from previous studies conducted in deeloping countries.^[**22, 33**]^ On the other hand, fear towards opiod related addiction was found significantly lower among respondents, and same type of attitudes were observed in significant studies^[28, 29, 33]^ but opposite attitudes were found in a large study conducted on the practicising physicians of Bangladesh.^[**33**]^

In recent years, multimodal analgesia has become increasingly widespread.^[**35**,**36**]^ Multimodal analgesia is achieved by combining different analgesics that act by different mechanisms and different sites in the nervous system, resulting in additive or synergistic analgesia with lowered adverse effects of sole administration of individual analgesics.^[**37**,**38**]^ This also refers to concurrent application of analgesic pharmacotherapy in combination with regional analgesia.^[**39**]^ It captures the effectiveness of individual agents in optimal dosages that maximize efficacy and attempts to minimize side effects. The advantages of this type of analgesia includes opiate-sparing effects as well as the goal of accomplishment of a more effective pain management though involving both central and peripheral anti-nociceptive mechanisms. Evidence suggests that using multimodal strategies provides better postoperative analgesia^[**40, 41**]^ and this approach can be used for day cases as well as for inpatient surgery.^[**42, 43**]^ So, multimodal analgesia significantly reduce postoperative hospital stay without increasing morbidity and mortality, in turn, decreasing patient’s costs and also increasing patient satisfaction.^[**44, 45**]^ The use of peripheral regional anesthetic techniques have been shown to be effective as a component of multimodal analgesia for management of postoperative pain associated with a number of surgical procedures^[**42, 43**]^.Although most of the students showed positive attitudes towards regional techniques, still considering multimodal analgesia as an economic burden to patients.This type of attitudes could be a barrier in future to appraise and implement evidence-based, procedure-specific multimodal analgesic protocolsin clinical practice. Overall, perception and attitudes of undergraduate medical students were found better in comparison to the studies conducted on medical students, interns and physicians of developing countries^[22, 33, 46]^ but there is still lack of knowledge in certin arenas which need to be addressed for achieving optimal management of postoperative pain.^[47, 48]^

## Conclusion

Majority of the patients who undergo surgical procedures experience acute postoperative pain due to inadequate management. Many postoperative pain management strategies are available to reduce postoperative pain. Current research found out that although most of the future physisians have well perception and attitudes towards postoperative pain management, still there were some sorts of confusion about opioids and multimodal analgesia. Incoorporation of pain education in undergratuate medical education might be fruitful for achieving optimal patient care.

## Data Availability

Data available on request

## ACKNOWLEDGEMENT(S)

The authors gratefully acknowledge the contributions of 4^th^ year medical students of studied medical colleges for their participation in this research.

